# Urinary microRNA Profiles Discriminate Bladder Cancer from Healthy Individuals: A Pilot Study Using qPCR Across Urine Fractions

**DOI:** 10.1101/2025.08.19.25333993

**Authors:** Cesar Augusto B. Duarte, Kamilla Leitão, Silvia Regina Hokazono, André Eduardo Varaschin, Sueli Massumi Nakatani

## Abstract

The development of reliable non-invasive diagnostic tools for bladder cancer remains a significant challenge. We hypothesized that the combined quantification of selected urinary microRNAs could differentiate bladder cancer cases from healthy controls. Using qPCR, we analyzed five candidate microRNAs (miR-21-5p, miR-141-3p, miR-210-3p, miR-200a-3p, and miR-103-3p) across three urine fractions: whole urine, pellet, and supernatant from a small cohort (n = 9). Although individual gene expression differences between cases and controls were not statistically significant (Wilcoxon p > 0.05), multivariate clustering using Mclust modeling and PCA showed clear separation, especially in the whole urine fraction. These results suggest that combined miRNA expression profiling, rather than individual markers, may provide diagnostic value for bladder cancer. Even though further evaluation in a larger population is necessary, our findings indicate that a cost-effective and easily implementable non-invasive diagnostic approach for bladder cancer may be within reach.

## 1. Introduction

Bladder cancer is among the most prevalent malignancies globally[1]. While cystoscopy remains the diagnostic gold standard, it is invasive, uncomfortable, and costly. There is increasing interest in identifying circulating and urinary microRNAs (miRNAs) as non-invasive biomarkers due to their remarkable stability in biofluids and potential disease specificity. Despite promising findings, the translation of miRNA-based diagnostics into clinical practice has been hindered by methodological inconsistencies.

A few reviews have brought light to the theme [2,3,4]. In a comprehensive and well-elaborated review, Grimaldi et al. highlighted the lack of standardization across critical pre-analytical and analytical steps, including the choice of urine fraction (e.g., whole urine, pellet, or supernatant), microRNA extraction methods, miRNA quantification techniques, and—importantly—the selection of reference genes for normalization. These sources of variability likely account for the wide array of proposed miRNA signatures for bladder cancer, many of which show inconsistent performance and lack convergence across studies. Consequently, although several candidate panels have demonstrated diagnostic promise, none have yet gained widespread adoption as robust tools for routine disease detection or monitoring.

In this pilot study, we sought to evaluate whether specific urinary miRNAs—either individually or in combination—can distinguish bladder cancer cases from controls across different urine fractions. Our approach emphasizes consistency in pre-analytical processing and normalization, aiming to contribute to the refinement of miRNA-based biomarker strategies. Importantly, by focusing on simple and accessible urine processing techniques and using qPCR-based quantification, we explored the feasibility of a cost-effective, easily implementable, and non-invasive diagnostic approach for bladder cancer that could ultimately evolve into a practical clinical tool.

## 2. Materials and Methods

### 2.1 Sample Collection & Design

Urine samples were collected from 5 bladder cancer patients and 4 healthy individuals. For the pellet fraction, only 4 cancer samples were analyzed due to sample availability.

### 2.2 miRNA Extraction & qPCR

Urinary fractions (whole urine, pellet, and supernatant) were processed using the miRNeasy Serum/Plasma Advanced Kit (Qiagen). Reverse transcription and qPCR were performed with the miRCURY LNA miRNA PCR Starter Kit (Qiagen) on a Roche LightCycler 480. For pellet and supernatant, one step centrifugation was performed, 3,000 rpm for 10 minutes.

### 2.3 Endogenous Control Selection

We evaluated endogenous control candidates based on expression stability (Ct < 32, CV < 0.15). miR-200a-3p exhibited the lowest CV and was used for normalization across all fractions.

### 2.4 Data Processing

Ct values were normalized by the selected endogenous control (ΔCt), followed by ΔΔCt transformation for case-control comparisons. Wilcoxon tests were performed for each miRNA.

### 2.5 Clustering and ROC Analysis

Mclust clustering was applied to ΔΔCt matrices (G = 2). Posterior probabilities were used to compute ROC curves via the “pROC” package.

2.6 This project received approval from the Genoprimer Diagnostico Molecular Research Ethics Board (approval n° 012).

## 3. Results

### 3.1 Ct Distributions and Endogenous Control Evaluation

Raw Ct values for five candidate microRNAs—hsa-miR-21, hsa-miR-141, hsa-miR-210, hsa-miR-200a, and hsa-miR-103a—were obtained from urine samples across three fractions: pellet, whole urine, and supernatant. These values are presented in Tables S1–3 (supplementary). Among the tested microRNAs, miR-200a-3p displayed the most stable expression profile across samples coefficient of variation (CV) ranging from 4.3% to 9.8% and was therefore selected as the endogenous control for normalization in subsequent analyses.

### 3.2 Statistical Comparison of Gene Expression

Normalized expression values were compared between cases and controls using the Wilcoxon rank-sum test. Although no microRNA reached conventional statistical significance (p > 0.05), miR-141 and miR-103 demonstrated trends toward differential expression (Table 1). This lack of statistical significance may be attributed to the limited sample size in this pilot study, underscoring the need for validation in larger, independent cohorts.

**Table 1.**
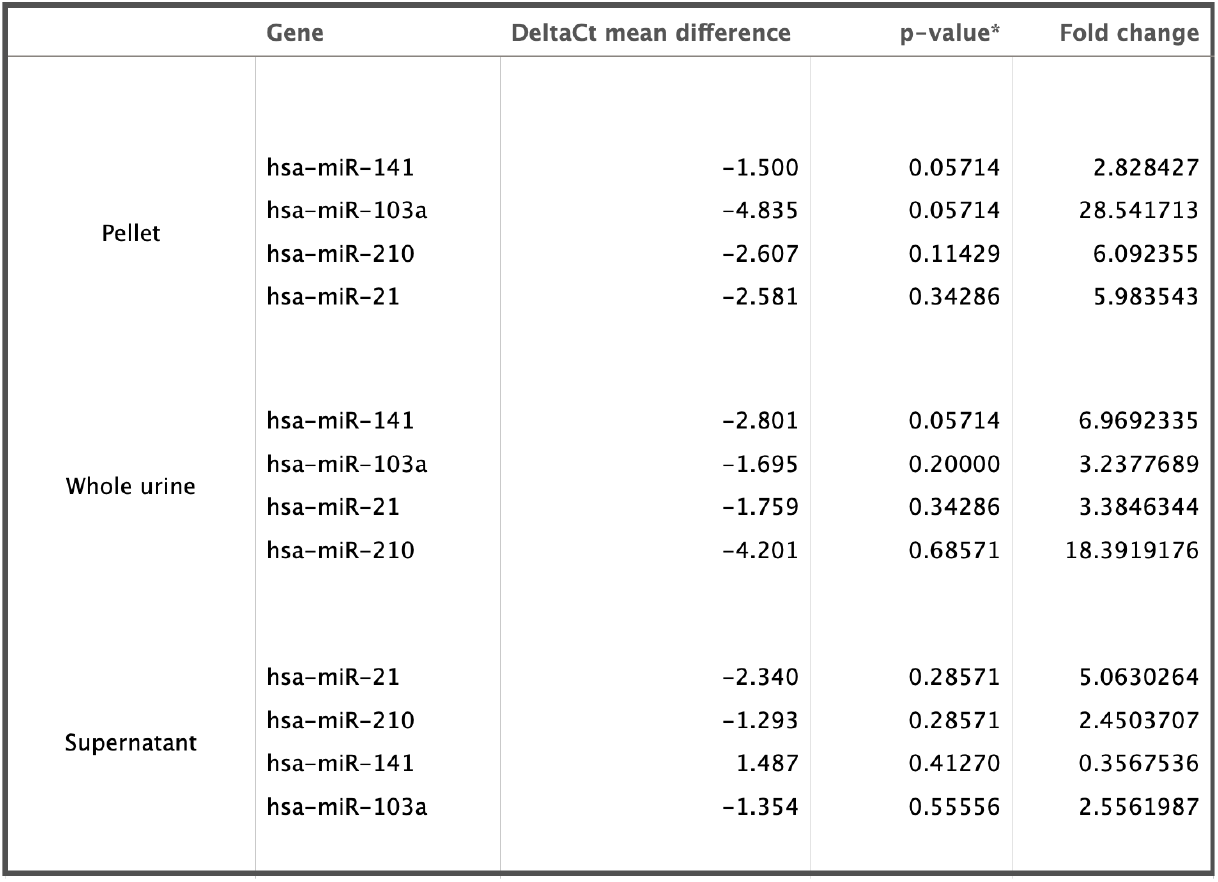
Differential expression analysis of urinary microRNAs between bladder cancer cases and controls.

Interestingly, all evaluated microRNAs exhibited consistently higher expression in bladder cancer samples compared to normal controls across the different urine fractions—with the exception of miR-141 in the supernatant, where its expression was lower in cases. The reason for this discrepancy is uncertain and may reflect technical variability. Notably, conflicting expression patterns for miR-141 have been reported in the literature, with some studies demonstrating upregulation [5] and others reporting downregulation [6] in bladder cancer. A similar pattern has been observed with another microRNA, miR-423-5p, which was found to be upregulated in whole urine [7] but downregulated in the supernatant fraction [8]. These discrepancies highlight the inherent variability—and at times, fragility—of miRNA-based biomarker strategies, reinforcing the importance of methodological standardization and validation in larger, well-characterized cohorts.

### 3.3 Mclust Clustering & ROC AUC

We applied unsupervised Mclust clustering to normalized data from each fraction, followed by classification performance assessment using ROC curve analysis:

Whole Urine: All cases clustered distinctly from controls. AUC = 1.0

Pellet: Two cancer samples were misclassified. AUC = 0.625

Supernatant: Cases and controls were intermixed. AUC = 0.70

### 3.4 PCA Analysis

PCA did not fully separate cases from controls in any fraction, though groupings were partially preserved, Fig. 1. Clustering via cosine similarity of sample vectors (angles between ΔΔCt vectors) yielded identical groupings to PCA in some cases, Fig.2, confirming reproducibility.

**FIG 1.**
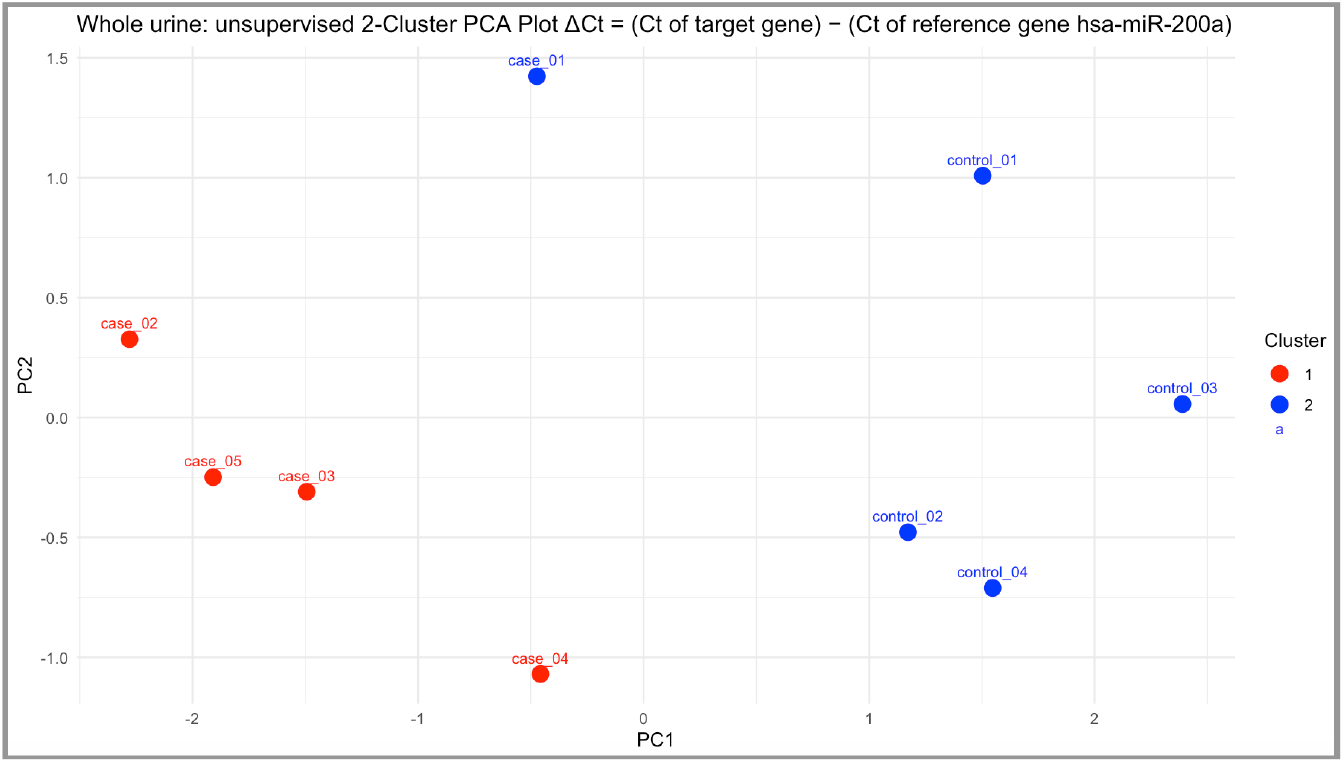
Principal component analysis (PCA) of urinary microRNA expression in bladder cancer cases and controls.

**FIG 2.**
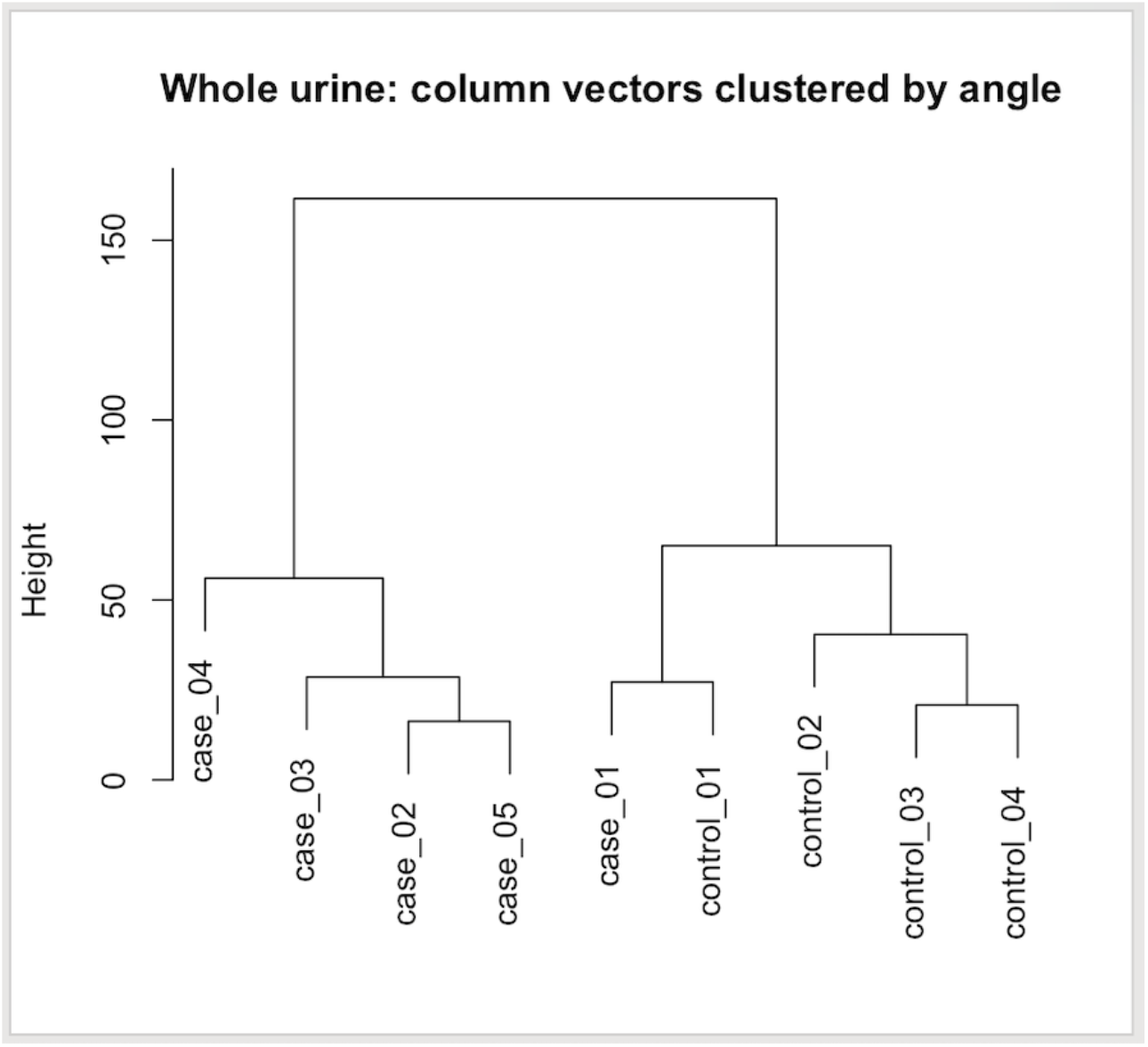
Clustering of urinary microRNA expression using cosine similarity of ΔΔCt vectors.

## 4. Discussion

Across all three urine fractions—pellet, whole urine, and supernatant—the expression patterns of the evaluated microRNAs showed a high degree of consistency. Most notably, all miRNAs except miR-141 in the supernatant displayed higher expression in bladder cancer cases than in controls, as reflected by negative ΔCt values and fold changes greater than 1. This pattern was especially prominent for miR-103a and miR-141 in the pellet and whole urine fractions, where fold changes were substantial (up to ∼28.5 for miR-103a). Such concordance in direction and magnitude of change across biologically distinct urine fractions suggests that these microRNAs may carry a genuine signal related to disease status, despite the lack of statistical significance in this small cohort.

Despite the lack of statistical significance on a per-miRNA basis, combining expression profiles via Mclust enabled separation of cases and controls—especially in whole urine. This suggests that composite expression patterns, rather than individual biomarkers, hold greater diagnostic promise. Endogenous control choice proved critical; miR-200a-3p consistently outperformed the provided candidate control miR-103a in terms of stability across our samples. It is worth noting, however, that the commercial qPCR system employed (miRCURY LNA miRNA PCR Starter Kit, Qiagen) was used in an off-label manner, as it is officially validated for use in serum, plasma, and urine exosomes—but not in whole urine or its individual fractions, which were the focus of our study.

The role of normalization remains a central challenge in miRNA-based diagnostics. In this study, miR-103 was evaluated as a candidate reference gene, as recommended by the commercial qPCR panel used. While miR-103 has been proposed in previous studies—particularly in combination with RNU48 [9], it did not meet the stability criteria adopted in our analysis and was therefore not selected for normalization. Interestingly, our data suggested that miR-103 may be upregulated in bladder cancer urine samples, in line with prior reports documenting its elevated expression in such patients [10]. This observation highlights a broader issue in the field: the potential for certain reference candidates to exhibit disease-associated variability, thus compromising their utility in normalization. These findings reinforce the need for the systematic identification and validation of robust, disease-invariant reference genes, specifically tailored to urinary miRNA profiling in bladder cancer. Without such rigor, the accuracy and reproducibility of biomarker-based diagnostics remain vulnerable to confounding influences.

Limitations in this present study include the small sample size (N = 9), which may inflate AUC due to overfitting. Further validation in larger, independent cohorts is essential. Nevertheless, this pilot study highlights the value of multivariate models and careful normalization in urinary miRNA biomarker research. Although our study focuses exclusively on the diagnostic potential of microRNAs for bladder cancer, it is noteworthy that the combined assessment of multiple biomarker types has also been explored. For instance, Eissa et al. [11] achieved high diagnostic accuracy by simultaneously measuring FOSB mRNA, RCAN1 mRNA, and the lncRNA miR-497-HG, in addition to miR-324-5p and miR-4738-3p.

## 5. Conclusion

Combined urinary miRNA expression profiling may represent a promising non-invasive strategy for bladder cancer detection. Among the urine fractions evaluated, whole urine appears to offer a practical advantage due to its simpler handling and reduced susceptibility to variation from centrifugation protocols—facilitating more consistent standardization across studies. Larger patient cohorts are needed to validate the diagnostic potential of the combined microRNAs tested in this study. Additionally, the use of sample vector clustering based on ΔΔCt angles showed trends similar to PCA and warrants further exploration as a potential modeling approach. Finally, integrating microRNAs with other biomarker classes may yield more robust diagnostic performance than microRNAs alone.

## Data Availability

All data produced in the present study are available upon reasonable request to the authors

