## Supplemental Tables for "Urinary microRNA Profiles Discriminate Bladder Cancer from Healthy Individuals: A Pilot Study Using qPCR Across Urine Fractions"

Table S1. Ct (cycle threshold) values of microRNAs from urine pellet fractions

| microRNA | case_01 | case_02 | case_03 | case_04 | control_01 | control_02 | control_03 | control_04 |
| --- | --- | --- | --- | --- | --- | --- | --- | --- |
| hsa-miR-21 | 18.86 | 21.18 | 25.11 | 20.64 | 28.58 | 26.46 | 25.41 | 23.78 |
| hsa-miR-141 | 24.58 | 23.43 | 27.43 | 28.42 | 33.20 | 29.80 | 28.50 | 26.48 |
| hsa-miR-210 | 23.05 | 23.53 | 30.01 | 28.19 | 32.52 | 33.22 | 30.14 | 27.45 |
| hsa-miR-200a | 27.07 | 26.88 | 29.42 | 31.73 | 34.88 | 31.14 | 28.68 | 28.53 |
| hsa-miR-103a | 21.76 | 25.13 | 23.44 | 23.44 | 30.10 | 30.52 | 30.31 | 30.31 |

Table S2. Ct values of microRNAs from whole urine fractions

| microRNA | case_01 | case_02 | case_03 | case_04 | case_05 | control_01 | control_02 | control_03 | control_04 |
| --- | --- | --- | --- | --- | --- | --- | --- | --- | --- |
| hsa-miR-21 | 28.22 | 20.48 | 28.05 | 31.11 | 25.10 | 26.91 | 30.93 | 32.23 | 31.07 |
| hsa-miR-141 | 28.27 | 25.69 | 28.99 | 31.28 | 30.99 | 30.36 | 30.37 | 33.99 | 31.78 |
| hsa-miR-210 | 39.79 | 26.57 | 28.55 | 28.55 | 28.55 | 39.71 | 32.47 | 39.81 | 32.48 |
| hsa-miR-200a | 31.63 | 28.40 | 32.46 | 32.04 | 32.53 | 29.81 | 30.76 | 31.71 | 30.76 |
| hsa-miR-103a | 30.30 | 25.77 | 31.27 | 30.30 | 30.30 | 30.46 | 31.46 | 32.47 | 31.46 |

Table S3. Ct values of microRNAs from urine supernatant fractions

| microRNA | case_01 | case_02 | case_03 | case_04 | case_05 | control_01 | control_02 | control_03 | control_04 |
| --- | --- | --- | --- | --- | --- | --- | --- | --- | --- |
| hsa-miR-21 | 29.47 | 23.34 | 29.63 | 32.96 | 30.22 | 30.69 | 31.96 | 35.24 | 34.26 |
| hsa-miR-141 | 34.34 | 26.89 | 30.98 | 34.93 | 35.49 | 32.38 | 32.19 | 31.97 | 33.90 |
| hsa-miR-210 | 32.90 | 29.73 | 33.56 | 35.4200 | 32.90 | 35.77 | 35.77 | 35.77 | 35.77 |
| hsa-miR-200a | 33.90 | 28.84 | 35.24 | 32.6600 | 32.66 | 33.32 | 34.42 | 34.23 | 34.96 |
| hsa-miR-103a | 34.61 | 29.28 | 40.72 | 34.6225 | 33.88 | 37.55 | 34.23 | 37.55 | 40.87 |
